# Prevalence of Occupation Associated with Increased Mobility During COVID-19 Pandemic

**DOI:** 10.1101/2020.12.11.20245357

**Authors:** Enbal Shacham, Stephen Scroggins, Matthew Ellis, Alexander Garza

## Abstract

**Objective:** Identifying geographic-level prevalence of occupations associated with mobility during local stay-at-home pandemic mandate.

**Methods:** A spatio-temporal ecological framework was applied to determine census-tracts that had significantly higher rates of occupations likely to be deemed essential: food-service, business and finance, healthcare support, and maintenance. Real-time mobility data was used to determine the average daily percent of residents not leaving their place of residence. Spatial regression models were constructed for each occupation proportion among census-tracts within a large urban area.

**Results:** After adjusting for demographics, results indicate census-tracts with higher proportion of food-service workers, healthcare support employees, and office administration staff are likely to have increased mobility.

**Conclusions:** Increased mobility among communities is likely to exacerbate COVID-19 mitigation efforts. This increase in mobility was also found associated with specific demographics suggesting it may be occurring among underserved and vulnerable populations. We find that prevalence of essential employment presents itself as a candidate for driving inequity in morbidity and mortality of COVID-19.

**Three-question Summary:**

1. Employees and workers deemed essential during the COVID-19 pandemic are likely to endure additional risk of infection due to community exposure. While preliminary reports are still quantifying this risk, we set out to examine if prevalence of specific occupations could be used to evaluate overall community-level risk based on stay-at-home mandate adherence.
2. Study results suggest that that not only are certain occupation geo-spatially associated with movement outside the home but are also associated with demographic characteristics likely to contribute to inequity of COVID-19 morbidity.
3. Often, nuanced inequities are lost in the larger data samples, being able to identify possible inequities from other sources such as prevalence of occupation among communities, remains an important and applicable alternative.

## Introduction

Inequity in incidence of morbidity and mortality related to the novel coronavirus (COVID-19) has become apparent across U.S. communities. Racial and ethnic minorities are experiencing higher rates of mortality, more rural areas are facing shortages in healthcare resources, and individuals living in poverty continue to present at hospitals in more advanced stages of COVID-19; decreasing the likelihood of positive outcomes.[1-3] Additionally, communities are confronting numerous economic and social challenges brought on by the pandemic; reflected by the nearly 40 million jobs lost, and only a small proportion regained, since the start of the pandemic in the U.S.[4, 5] However, for individuals in frontline employment, or essential occupations, who leave the home for work, inequity and risk of COVID-19 is not yet fully understood. As U.S. mortality reaches over 200,000 as of October 2020, there is a need to identify risk more effectively among vulnerable populations.[6]

Throughout the U.S., state and local governments have implemented mitigation mandates to slow the spread of COVID-19.[7] Commonly and with varying degrees of efficacy, these efforts include face-covering requirements, limitations on business capacities, changes to hours of business operations, social-distancing, and recommendations to stay at home.[8] By the nature of viral transmission, staying home remains the most effective COVID-19 prevention method, though it also remains the most challenging for some. Among U.S. states, 42 have published guidelines and mandates of what may constitute an essential worker; an individual employed in an occupation required to be present at a work, largely unable to stay at home. The U.S. Department of Homeland Security defines an essential worker as someone with a range of responsibilities whose continued work is critical to infrastructure operations.[9] Historically, an essential worker has become synonymous with emergency personal: health workers, law enforcement, and those in safety occupations. However, recent research, has shown essential workforce during the COVID-19 pandemic is likely to include transportation workers, employees of restaurants and grocery stores, educators, among other seemingly unrelated occupations.[10] Among these newly-defined essential workers, the burden of community mobility and increased chance of COVID-19 exposure remains unclear both individually and ecologically.

Historically, there has been little understood about the impact that community mobility has had on the health of an individual. Previous location-anchored research focused on home or work address, yet the movement throughout an individual’s daily lives offers insights, in this case particularly to potential COVID-19 exposure. The purpose of this study was to determine the association among prevalence of occupations and work-related mobility framed within the current COVID-19 pandemic.

## Materials and Methods

This study employed an ecological design with observations at the census-tract level among two densely populated urban Midwest counties (n=305). The study period consisted of all days from March 16, 2020 to May 17, 2020; coinciding with seven days before a stay-at-home mandate for all 1.3 million approximate residents within census-tracts and fifty-six days during the stay at home mandate.

Two secondary data sources were utilized for this study. The 2018 5-year American Community Survey (ACS) was used to determine proportion of workers, aged 16 and older, in occupations of interest among each census-tract. These occupations included: 1) healthcare support, 2) food preparation and service, 3) building, ground maintenance 4) management, business, and finance, and 5) office administration support.[11] The ACS was also used to acquire socio-demographic variable per-census-tract that were likely to be associated with adverse health outcome; (1) proportion racial/ethnic minority, (2) proportion earning wages below the federal poverty level, (3) average number of people per household, (4) population density per-square mile, and (5) proportion of residents aged >64.

Mobility among census-tracts was estimated using aggregated and anonymized data and acquired as part of a COVID-19 research consortium managed by SafeGraph, LTD, a data management firm. This data was collected using mobile smart device pings collected throughout the day and used to estimate geo-global position. This large dataset contains an estimated 5-10% of the U.S. population. Residency of devices, used to establish proportion of sample leaving home, was calculated by determining the location the device during overnight hours of the day over the course of six weeks.[12] Additionally, this data has been leveraged for similar studies and surveillance, used by state and national organizations.[13]

Descriptive statistics were calculated for all variables. A series of Getis-Ord Gi∗ cluster analysis were applied among census-tracts for each occupation category to determine locations where they were significantly higher or lower.

Each occupational category was used as a primary predictor in a series of multivariate spatial regression models predicting the average proportion of census-tract residents adhering to the stay-at-home order. Each model was adjusted for socio-demographics described a priori as well as the average proportion of census-tract residents staying home seven days before the stay-at-home order as a baseline measurement of mobility. Each model was first constructed under ordinary-least square regression assumptions and a diagnostic cascade was used to detect if spatial dependence was present. Preliminary results indicated that adjusting for this dependency was necessary, thus spatial-lag models were determined to be the best fit.

Both descriptive and inferential analyses were completed using R 4.0 and GeoDa 1.14.[14, 15] Significance was determined at α of 0.05 and spatially detected using a pseudo p-value permutation. Spatial weights were applied using a queen contiguity matrix.

## Results

As shown in Table 1, prior to the stay-at-home policy, a daily average of 34.75% +4.92 of the sample did not leave home. During the eight weeks of the stay-at-home policy, a daily average of 41.59% +5.51 did not leave home. Average proportions of census-tract residents employed in the occupations of interest included: 15.86% +8.06 working in management, business, and finance, 12.37%+4.91 working in office administration support; 6.44% +4.24 working in food preparation and service; 4.25% +4.65 working in healthcare support; and 4.13% +3.68 working in building, ground maintenance.

**Table 1.** Characteristics of Census-tracts within City of St. Louis and St. Louis County (n=305)

|  |  |
| --- | --- |
|  | Mean % (SD) |
| Stay-at-Home Policy Adherence |  |
| Daily % not leaving home before policy<sup>a</sup> | 34.75 (4.92) |
| Daily % not leaving home during policy<sup>b</sup> | 41.59 (5.51) |
| Non-white population | 44.47 (33.42) |
| Employed but income below federal poverty level | 13.99 (11.11) |
| Residents within household | 2.36 (0.34) |
| Population density (mi<sup>2</sup>) | 4,317.36 (2,766.66) |
| Older adults, aged $\geq 65$ | 15.70 (5.83) |
| Occupation |  |
| Healthcare support | 4.25 (4.65) |
| Food preparation and service | 6.44 (4.24) |
| Building, ground maintenance | 4.13 (3.68) |
| Management, business, and finance | 15.86 (8.06) |
| Office administration support | 12.37 (4.91) |
| a. Calculated from daily observations from March 16, 2020 to March 22, 2020 |  |
| b. Calculated from daily observations from March 23, 2020 to May 17, 2020 |  |

Figure 1 details significant geographical clusters (p<0.05) where each occupation category was higher or lower than dispersion would expect. Among the 305 census-tracts, 50 (16.39%) were identified as having significantly higher proportion of healthcare support workers (Fig 1a); 33 (10.81%) as having significantly higher proportion food service and preparation workers (Fig 1b); 57 (18.68%) as having significantly higher building, ground maintenance workers (Fig 1c); 66 (21.64%) as having significantly higher proportion of business, management, and finance workers (Fig 1d); and 46 (15.08%) identified as clusters with higher proportion of office administration workers (Fig 1e).

**Figure 1.**
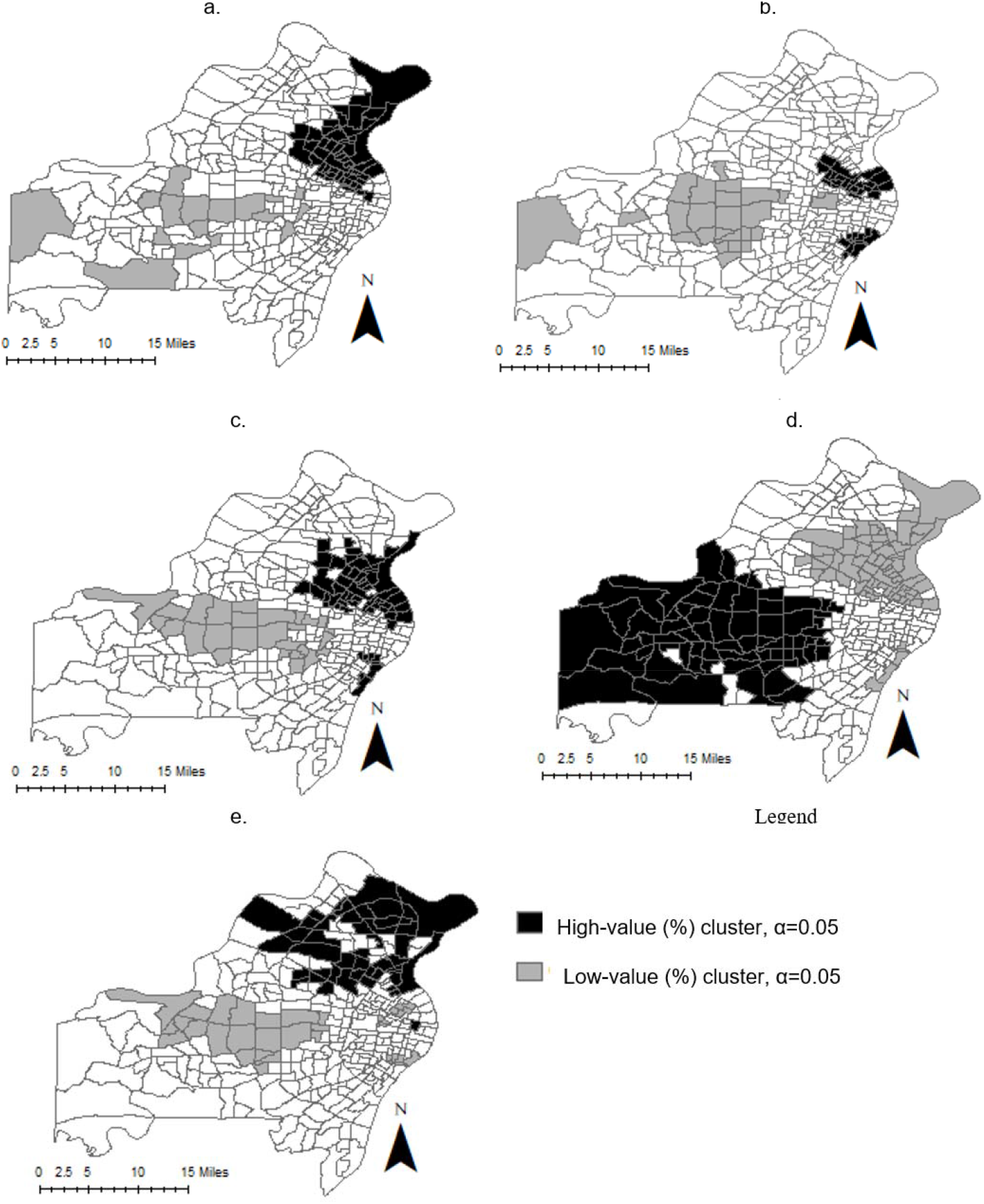
Significant Gi* clustering of censustracts (n=305) in St. Louis City and County according to proportion of selected occupation categories: (a). Proportion working in healthcare support, (b). Proportion working in food service and preparation, (c). Proportion working in building, ground maintenance, (d) Proportion working in management, business, or finance, (e). Proportion working in office administration support.

Table 2 details five spatial regression models, each adjusted for spatial lag, predicting proportion of residents staying home during stay-at-home policy implementation and adjusted for socio-demographics. Model 1 reveals that for every 10% increase in healthcare support workers, the proportion staying home decreases by 1% per-census-tracts (β =-0.1, p=0.020). Similarly, every 10% increase in food service and preparation workers results in a 1% decrease in persons staying home (model 2, β=-0.01, p=0.019) Model 5 shows the effects of office administrative workers per census-tract; a 16% increase results in a 1% decrease in persons staying home (β=0.06, p=0.042). Model 4 displays an opposite pattern; for every 12% increase in proportion of workers in management, business, or finance occupations the proportion of persons staying home increased by 1% (β=0.08, p=0.002). The occupation category of building, ground maintenance was found not to be a significant spatial predicator of COVID-19 mitigation compliance when adjusting for tract characteristics (model 3).

**Table 2.**
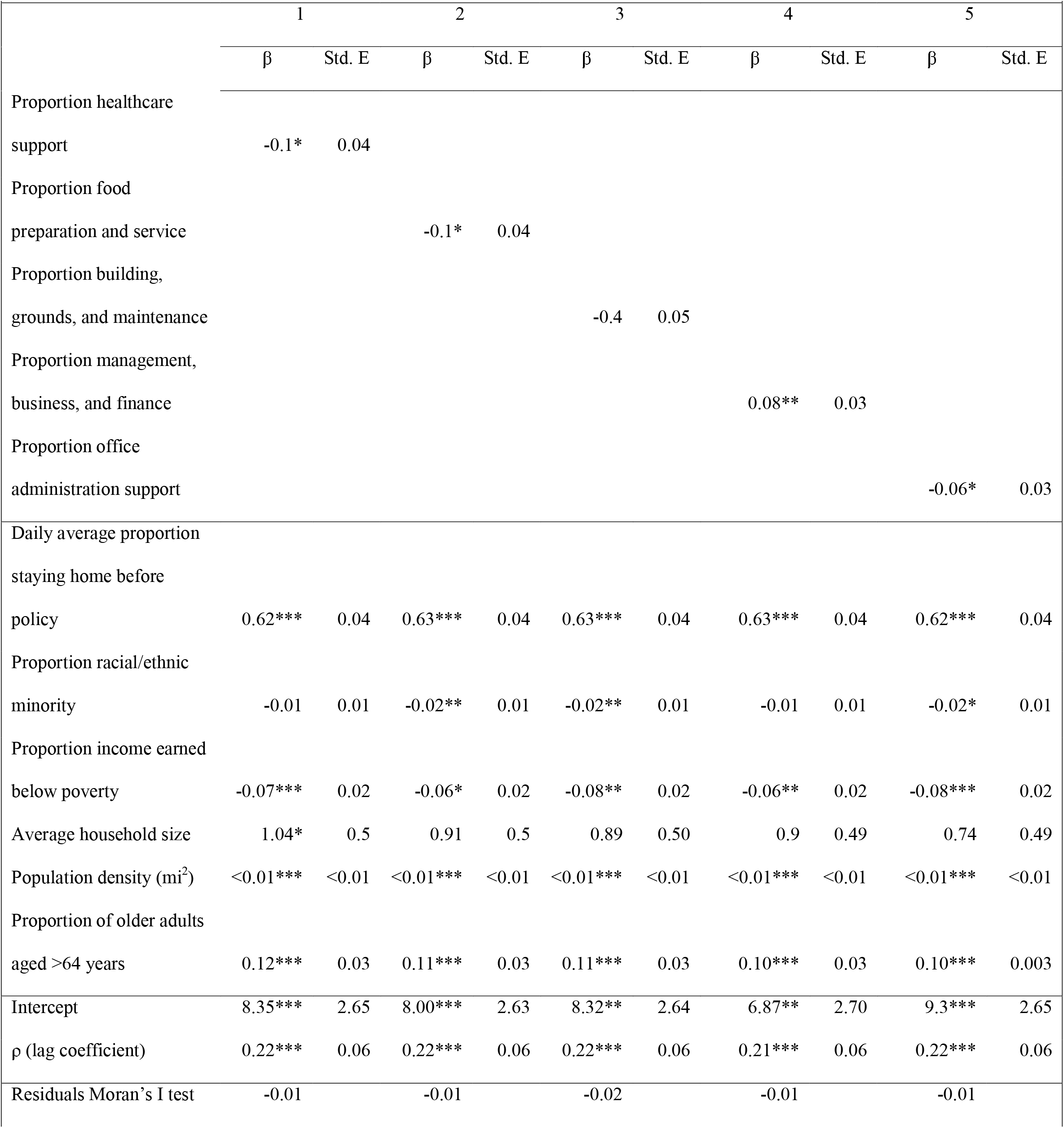
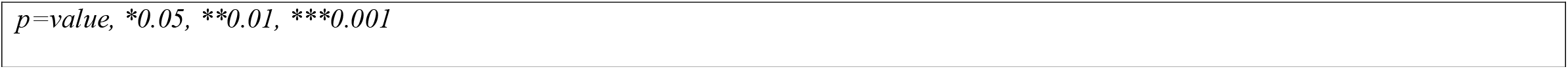
Spatial Regression Models Predicting Proportion Residents Adhering to Stay-at-Home Policy Implementation

|  | 1 |  | 2 |  | 3 |  | 4 |  | 5 |  |
| --- | --- | --- | --- | --- | --- | --- | --- | --- | --- | --- |
| | $\beta$ | Std. E | $\beta$ | Std. E | $\beta$ | Std. E | $\beta$ | Std. E | $\beta$ | Std. E |
| Proportion healthcare support | -0.1* | 0.04 |  |  |  |  |  |  |  |  |
| Proportion food preparation and service |  |  | -0.1* | 0.04 |  |  |  |  |  |  |
| Proportion building, grounds, and maintenance |  |  |  |  | -0.4 | 0.05 |  |  |  |  |
| Proportion management, business, and finance |  |  |  |  |  |  | 0.08** | 0.03 |  |  |
| Proportion office administration support |  |  |  |  |  |  |  |  | -0.06* | 0.03 |
| Daily average proportion staying home before policy | 0.62*** | 0.04 | 0.63*** | 0.04 | 0.63*** | 0.04 | 0.63*** | 0.04 | 0.62*** | 0.04 |
| Proportion racial/ethnic minority | -0.01 | 0.01 | -0.02** | 0.01 | -0.02** | 0.01 | -0.01 | 0.01 | -0.02* | 0.01 |
| Proportion income earned below poverty | -0.07*** | 0.02 | -0.06* | 0.02 | -0.08** | 0.02 | -0.06** | 0.02 | -0.08*** | 0.02 |
| Average household size | 1.04* | 0.5 | 0.91 | 0.5 | 0.89 | 0.50 | 0.9 | 0.49 | 0.74 | 0.49 |
| Population density (mi<sup>2</sup>) | <0.01*** | <0.01 | <0.01*** | <0.01 | <0.01*** | <0.01 | <0.01*** | <0.01 | <0.01*** | <0.01 |
| Proportion of older adults aged >64 years | 0.12*** | 0.03 | 0.11*** | 0.03 | 0.11*** | 0.03 | 0.10*** | 0.03 | 0.10*** | 0.003 |
| Intercept | 8.35*** | 2.65 | 8.00*** | 2.63 | 8.32** | 2.64 | 6.87** | 2.70 | 9.3*** | 2.65 |
| $\rho$ (lag coefficient) | 0.22*** | 0.06 | 0.22*** | 0.06 | 0.22*** | 0.06 | 0.21*** | 0.06 | 0.22*** | 0.06 |
| Residuals Moran's I test | -0.01 |  | -0.01 |  | -0.02 |  | -0.01 |  | -0.01 |  |

$p$ =value, \*0.05, \*\*0.01, \*\*\*0.001

Within regression models, several of the socio-demographic characteristics were determined to be significant predictors of populations staying home. Consistent across all models, as proportion of older adults and population density increased, proportion of census-tract residents staying home also increased. As proportion of those earning income below the poverty level increased, the proportion of tract residents staying home decreased significantly in each model. Average household size and proportion racial/ethnic minorities per-census-tract were also identified as significant model predictors, although inconsistently throughout models.

## Discussion

This study was able to identify relationships among prevalence of occupations and patterns of community mobility that have the potential increase risk for COVID-19 exposure among residents and communities. Areas where census-tract residents were proportionally less likely to remain at home were also likely to have more individuals employed in the fields of healthcare support, food preparation and service, and office administration support. In addition, our study results point to higher proportions of vulnerable populations sharing similar mobility patterns. These types of occupations identified within this study often require fewer credentials, pay substantially less, offer minimal flexibility, and are more likely to be among the limited employment opportunities available to racial and ethnic minorities and residents of low-income communities.[16, 17]

These findings highlight the accelerated impact of inequity related to COVID-19. While employment typically serves as a protective factor in many health outcomes, this pandemic has shown that types of employment and the varying context of essential employment change, leaving already vulnerable employees at increased risk for adverse health outcomes. Current literature supports this framework, finding that pandemic essential employment sites require ensuring proper protective equipment, sanitization, and safety protocols, yet, in many cases, are unavailable due the challenges in understanding the transmission and supply chain demands.[18]

Community mobility, in this case, provided insights to where and when individuals were home and work during a stay-at-home order. These methods and results highlight the utility of these types of data and how they can provide insights to patterns of behavior and health outcomes throughout communities. Other studies have more widely begun to use these methods and have great opportunity to the growth of our science about how neighborhoods impact our health. {Painter, 2020 #2595}

The pandemic has provided an opportunity for employers, employees, and the public to re-consider such positions now deemed “essential workers.” This will hopefully pave the way for acknowledging and addressing economic inequities that existed prior to the pandemic, but have gained greater visibility, not only in terms of employee safety, but employee value and risk. Efforts such as raising the federal minimum wage and improving the accessibility of healthcare coverage should now be included in conversations highlighting lessons learned from the current pandemic.

## Data Availability

Data for this study is owned and managed by Safegraph, LCC. Access to data was granted as part of a national COVID-19 Research Consortium.

## Acknowledgements

This work was supported by the Sinquefield Center for Applied Economic Research and the Geospatial Institute at Saint Louis University.

